# The Validity of the Parsley Symptom Index: an e-PROM designed for Telehealth

**DOI:** 10.1101/2022.04.15.22273917

**Authors:** Hants Williams, Sarah Steinberg, Kendall Leon, Catherine O’Shea, Robin Berzin, Heather Hagg

## Abstract

**Background / Purpose:** The Parsley Symptom Index (PSI) is a recently developed symptom assessment for adults with chronic disease in telehealth settings. The purpose of this study was to validate the PSI against the Self-Rated Health (SRH) item.

**Materials and Methods:** This prospective cohort study took place from January 15, 2021 to December 15, 2021 among a sample of 10,519 adult patients at Parsley Health, a subscription based holistic medical practice. The PSI and the SRH were completed by patients via an online portal. The association between the PSI and SRH was assessed via polyserial and polychoric correlations, while weighted kappa scores provided information related to agreement between the PSI and SRH.

**Results:** From 22,748 responses, there were moderate levels of association (polyserial *r*=0.51; polychoric *r*=0.52) and agreement (weighted □ = 0.46) between the PSI and SRH. In total 74.2% (16865) of responses between the PSI and SRH were relatively congruent while 36.2% (8229) were literally congruent.

**Conclusions:** The PSI demonstrates validity with the SRH for adults with chronic disease in a telehealth setting.

## Introduction

Providing telehealth options has become indispensable to healthcare delivery. Even before the COVID-19 pandemic fundamentally altered the healthcare landscape, claim lines for non-hospital-based clinicians-to-patient telehealth grew 1,393%^1^from 2014 to 2018.. Health crisis triaging during the COVID-19 pandemic further increased demand for telehealth care^2,3^, accelerating the transition from brick and mortar practice to the virtual interface. The pandemic spawned an entirely new telehealth industry, reducing access and cost barriers for patients, from the rural farmer to the busy urban professional^4 5.^

Having access to affordable care is especially important for the 60% of Americans that live with at least one chronic disease -this group spends 2-4 times more on healthcare than those without any chronic conditions^6^. Telehealth helps clinicians effectively manage chronic disease with increased opportunity to monitor treatments and quickly respond to patient concerns^7^, which reduces costs^8^ and hospitalizations^9^. Despite the fact that many people with chronic diseases are currently being treated via telehealth, there are limited tools specifically designed for virtual chronic care available to telehealth providers and clinics. Electronic patient-reported outcome measurement (e-PROMs) is one tool that serves as the first patient touchpoint in telehealth generally, and in particular can play a pivotal role in the clinical care of patients with chronic conditions. Completing e-PROMs allows patients to reflect on their own health, boosts patient-clinician communication, and empowers patients to steer their own healthcare journey^10^.

As part of a larger effort to leverage new tools like e-PROMs to make the telehealth experience engaging and effective for patients with chronic diseases, a research team at Parsley Health–a subscription based holistic medicine practice– built the Parsley Symptom Index (PSI). The PSI is a 45-item e-PROM designed specifically for use in telehealth settings to function as a Review of Systems (ROS). When used strategically, a patient reported outcome driven approach can shift a ROS to a cooperative dialogue between patients and clinicians^11^. Like a ROS, the PSI focuses on bodily domains, and the most commonly reported symptoms associated with chronic conditions for each domain. As a digital first e-PROM, we built the PSI to provide immediate feedback to patients, producing data that is seamlessly adopted into the standard clinical workflow and provides the scaffold for an effective patient-clinician conversation^12^. To our knowledge the PSI is the only existing short form e-PROM developed with preliminary validation for use within a 100% telehealth setting with chronic disease patients^13^.

In an initial feasibility and acceptability study that accessed construct and face validity, the PSI was deployed, completed, and found helpful to both patients and clinicians^13^. Having previously described the item generation, accessibility and interpretability in a population receiving longitudinal care, the aim of this study is to continue validation of the PSI by comparing it against the Self Rated Health score (SRH), a single-item question that has been successfully used in prior research to test construct validity of patient perceived health^14,15,16^.

## Materials and Methods

### Study Design

This retrospective cohort study took place at Parsley Health from January 15, 2021 to December 15, 2021 among a sample of 10,519 adult patients. Patients completed the PSI and the SRH via an online portal. This study used patient-reported survey data that was recorded in such a manner that participants were unidentifiable to the researchers. The Institutional Review Board at Stony Brook University considered this study exempt from 45 Code of Federal Regulations requirements^17^.

### Study Setting and Population

Parsley Health is a subscription-based membership model for delivering primary care and proactive chronic disease management through a holistic medicine lens. Patients receive care from Parsley Health clinicians and health coaches in-person and virtually, with additional access to their care team via email and an online portal. Prior to the COVID-19 pandemic Parsley Health provided telehealth across 10 states in addition to in-person care. In the COVID-19 pandemic Parsley Health further increased their telehealth availability to over 45 states. Inclusion criteria for this study were Parsley Health patients that had (1) an active subscription membership plan between January 15, 2021 and December 15, 2021, and (2) a minimum of one clinical encounter within their membership period. Exclusion criteria were (1) severe psychiatric disorders (particularly psychosis and depression requiring a change in treatment in the last 30 days); (2) under the age of 18; or (3) unable to speak or read English.

### Parsley Symptoin Index (PSI)

The PSI is a 45-item, ROS-style patient reported outcome measurement (PROM) tool designed to capture chronic disease symptoms^13^. Items are grouped into nine systems, with each containing four to seven items per group that are ranked on a scale from 0 (asymptomatic) to 10 (extremely symptomatic). A total score is calculated with the following four cut-off ranges: 0-24, 25-43, 44-71, and greater than 71. The following terminology for these ranges are “well” (0-24), “symptomatic” (25-43), “very symptomatic” (44-71), and “sick” (71+). Upon completing the PSI, patients can immediately view their PSI score in graphical format and compare it to past responses, stratified by body systems.

### Self-Rated Health (SRH) Item

To support validation of the PSI, a mandatory SRH item was administered with the PSI to ensure a SRH was completed for every PSI. The SRH is a single question, with a 5-item likert scale answer that reads: “In general, would you say that your health is excellent, very good, good, fair, or poor?” The SRH is validated and is commonly used to demonstrate construct validity of PROMs^14 15 16^, and allows the clinician to do a quick global assessment of patient perceived well-being.

### Integration Into Clinical Workflow

After patients scheduled a visit, they were instructed to log into an online patient portal and complete the PSI 24-48 hours before each clinical visit. Initial visits were rescheduled if all forms were not completed, but follow-up visits were not postponed for an incomplete PSI. For follow-up visits, patients who hadn’t completed the PSI received an automated reminder 48 hours before the clinical visit. If the PSI was not completed after the automated prompt, another prompt was sent from the clinician or clinical operations coordinator.

When clinicians prepared for an online visit, they used a standardized note template within the electronic health record to pull the most recent PSI score into the visit note. The PSI design allowed for the results to be immediately usable: once a PSI was completed, patients received instant feedback, and clinicians could quickly import the data into the note to prepare for the patient visit. With the PSI template integrated into the beginning of the encounter note, clinicians were subtly prompted to use the PSI to discuss patient reported symptoms and provide positive feedback to the patient for completing the PSI.

During the telehealth patient visit, the PSI score was used as a touchpoint for the patient– clinicians discussion. As the PSI was previously completed, clinicians were able to ask targeted questions about symptoms and had more time to focus on burden and distribution of illness. The longitudinal PSI graph further deepened the provider’s ability to identify triggers and mediators that influenced disease trajectory over time.

### Association Analysis

To test the hypothesis that the SRH item would correlate with the PSI, two measures of association were calculated. First a polyserial correlation was performed on the raw continuous score of the PSI (range 0-500) with the ordinal SRH categories (Excellent, Very Good, Good, Fair, Poor). Next, the PSI’s responses were scored and translated into ordinal categories (1=Great, 2=Good, 3=Average, 4=Fair, 5=Poor) to compare directly with the SRH categories and generate polychoric correlation coefficients^19^. This second analysis provided an alternative view for when the PSI is interpreted as ordinal instead of continuous.

### Agreement Analysis

To determine agreement, weighted kappa (quadratic) scores incorporated information about the distance between the transformed ordinal PSI and SRH ratings: ratings that were one category apart counted as “less disagreement” than a pair of ratings two categories apart. The weighted Kappa method partially contributes to responses that are “near” the rating category–for example, “very good” to “excellent” is categorically closer than “excellent” to “poor”. To interpret kappa, the following guidelines are used to suggest agreement^20,21,,22,23^.

- 0 = agreement equivalent to chance
- 0.10–0.20 = slight agreement
- 0.21–0.40 = fair agreement
- 0.41–0.60 = moderate agreement
- 0.61–0.80 = substantial agreement
- 0.81–0.99 = near-perfect agreement
- = perfect agreement

In addition, a binary interpretation of agreement results as ‘literally congruent’ or ‘relatively congruent’ were calculated. If the PSI and the SRH were an exact match (e.g., both scored as ‘Very Good’, or both ‘Poor’) the congruence type was scored as literal, while if an individual’s responses to the PSI and SRH were not an exact match but consistent in terms of their position as either good (‘Excellent’, ‘Very Good’, ‘Good’) or bad health (‘Fair’, ‘Poor’) the congruence type was scored as relative^24^.

### Data Analysis Software

All analyses were carried out in SAS v9.4.^25^

## Results

There were a total of 22,732 observations from 10,519 unique patients from January 15 2021 to December 15 2021. Only completed sociodemographic data for patients are represented in Table 1. Race and gender identity data are not complete for the entire sample, and were added in late January 2020 for new members. Missing data for member race and gender that had registered prior to January 2021 are still being retroactively collected by staff. Data describing race/ethnicity and gender identity refer to the segment of the population for which that data is complete (N=8,034 for gender, N=7,919 for race/ethnicity).

**Table 1:** Patient Descriptives.

|  | Level | Overall |
| --- | --- | --- |
| **n** |  | 10531 |
| **Biological Sex (%)** | Female | 9092 (86.3) |
|  | Male | 1351 (12.8) |
|  | Other | 88 (0.8) |
| **Gender Identity (%) \*** | Woman | 6942 (86.3) |
|  | Man | 1011 (12.6) |
|  | Non-binary | 33 (0.4) |
|  | Female to Male | 13 (0.2) |
|  | Male to Female | 11 (0.1) |
|  | Non-binary Other | 13 (0.2) |
|  | Transgender | 6 (0.1) |
|  | Gender Queer | 13 (0.2) |
| **Race (%) \*** | White | 6104 (75.9) |
|  | American Indian or Alaskan Native | 27 (0.3) |
|  | Asian | 508 (6.3) |
|  | Black or African American | 560 (7.0) |
|  | Native Hawaiian or other Pacific Islander | 23 (0.3) |
|  | Other | 690 (8.6) |
|  | Prefer not to say | 130 (1.6) |
| **Age Group (%)** | 18-24 | 459 (4.4) |
|  | 25-34 | 3931 (37.3) |
|  | 35-44 | 3346 (31.8) |
|  | 45-54 | 1604 (15.2) |
|  | 55-64 | 783 (7.4) |
|  | 65-74 | 326 (3.1) |
|  | 75-84 | 74 (0.7) |
|  | 85+ | 8 (0.1) |
| **Number Medical Visits (mean [SD])** |  | 3.07 (3.11) |
| **Number Health Coach Visits (mean [SD])** |  | 2.30 (2.81) |
| **Total Membership Duration** | 0 to 1 year | 7361 (69.9) |
|  | 1 to 2 years | 1755 (16.7) |
|  | 3 or more years | 1415 (13.4) |
| **Most Frequent ICD Codes** | Abdominal distension (gaseous) | 3300 (31.3) |
|  | Other fatigue | 3244 (30.8) |
|  | Anxiety disorder, unspecified | 2554 (24.2) |
|  | Irritable bowel syndrome with diarrhea | 1826 (17.3) |
|  | Constipation, unspecified | 1626 (15.4) |
|  | Insomnia, unspecified | 1105 (10.4) |
|  | Hypothalamic dysfunction, not elsewhere classified | 1079 (10.2) |

The distribution of responses for each scale item was skewed toward the positive (Table 2). 12.5% (2834) of respondents reported their health as *Excellent* for the PSI and 3.6% (817) for the SRI, while 22.9% (5207) and 38.7% (8794) rated their health as *Very good* or *Good* for the PSI and 25.3% (5759) and 42.8% (9734) for the SRI. Fewer than 25% (5897) rated their health to be *Fair* or *Poor* on the PSI and 27% (6422) for the SRI.

**Table 2:**
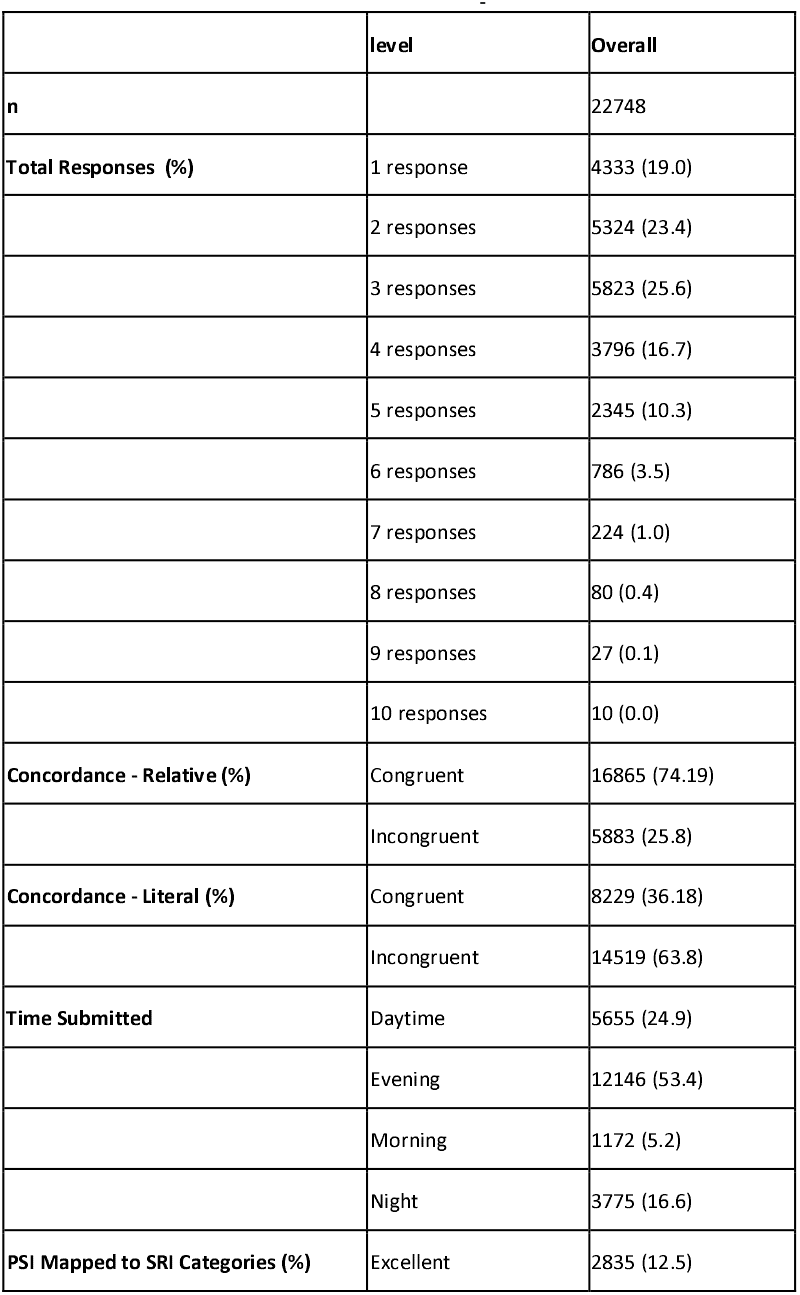
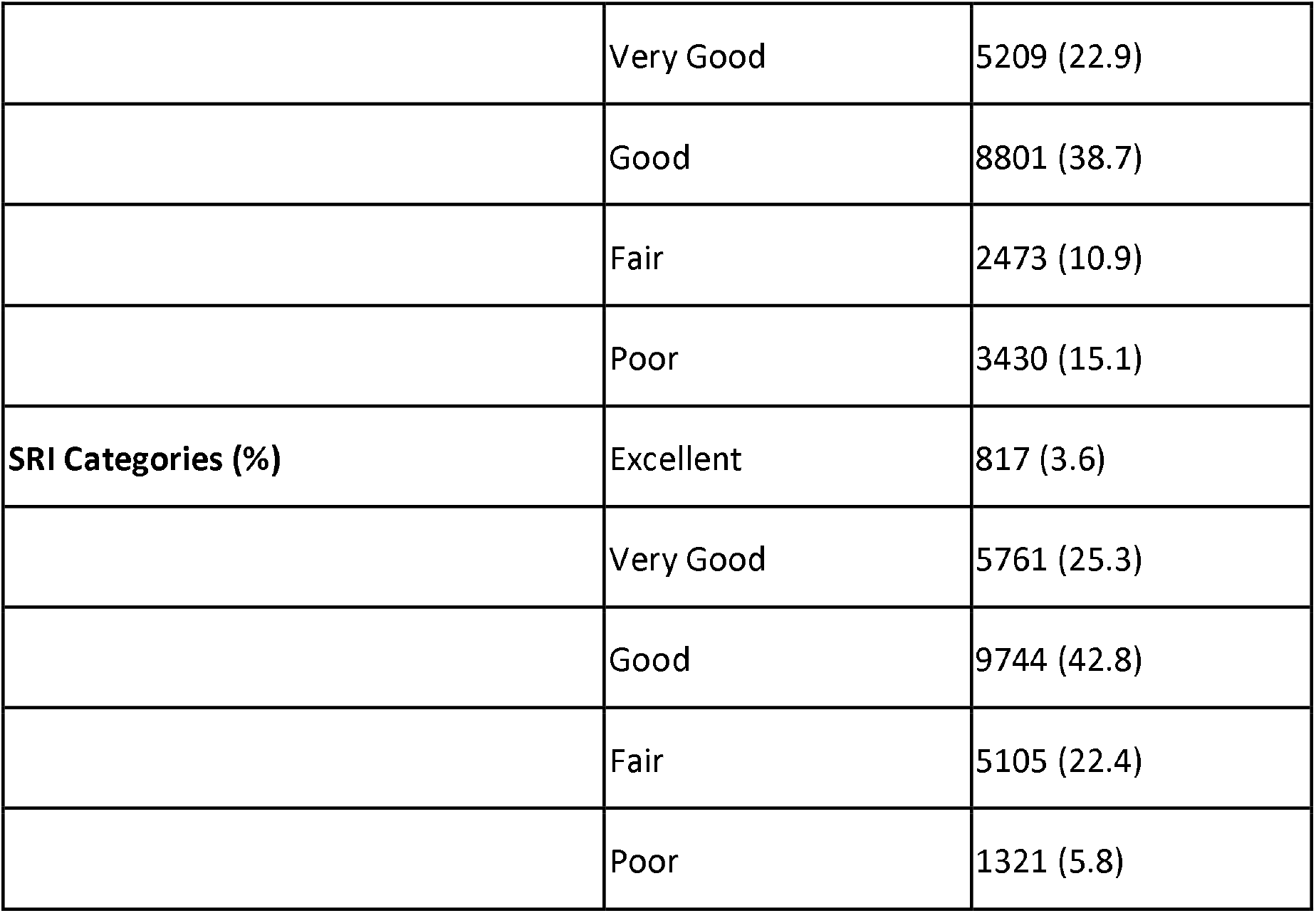
PSI and SRH Descriptives.

|  | level | Overall |
| --- | --- | --- |
| n |  | 22748 |
| Total Responses (%) | 1 response | 4333 (19.0) |
|  | 2 responses | 5324 (23.4) |
|  | 3 responses | 5823 (25.6) |
|  | 4 responses | 3796 (16.7) |
|  | 5 responses | 2345 (10.3) |
|  | 6 responses | 786 (3.5) |
|  | 7 responses | 224 (1.0) |
|  | 8 responses | 80 (0.4) |
|  | 9 responses | 27 (0.1) |
|  | 10 responses | 10 (0.0) |
| Concordance - Relative (%) | Congruent | 16865 (74.19) |
|  | Incongruent | 5883 (25.8) |
| Concordance - Literal (%) | Congruent | 8229 (36.18) |
|  | Incongruent | 14519 (63.8) |
| Time Submitted | Daytime | 5655 (24.9) |
|  | Evening | 12146 (53.4) |
|  | Morning | 1172 (5.2) |
|  | Night | 3775 (16.6) |
| PSI Mapped to SRI Categories (%) | Excellent | 2835 (12.5) |
|  | Very Good | 5209 (22.9) |
|  | Good | 8801 (38.7) |
|  | Fair | 2473 (10.9) |
|  | Poor | 3430 (15.1) |
| **SRI Categories (%)** | Excellent | 817 (3.6) |
|  | Very Good | 5761 (25.3) |
|  | Good | 9744 (42.8) |
|  | Fair | 5105 (22.4) |
|  | Poor | 1321 (5.8) |

The polyserial correlation between raw PSI scores and the SRH was *r* = 0.51, suggesting moderate association. When treating PSI scores as ordinal (transformed to SRH scale) the polychoric correlation coefficient was nearly identical at *r* = .52, also suggesting moderate association. The weighted kappa coefficient between the transformed PSI and SRH was .46 suggesting moderate agreement (Table 3). The agreement analysis shows approximately 74.2% (16,865) relative congruence and 36.2% (8,229) literal congruence across all observations.

**Table 3:** Association and Agreement.

|  | Response count (%) | Polyserial<br>Correlation | Polychoric<br>Correlation | Relative<br>Concordance | Literal<br>Concordance | Weighted<br>Kappa | Maximum<br>Kappa |
| --- | --- | --- | --- | --- | --- | --- | --- |
| **Total** | 22732 (100%) | 0.517 | 0.522 | 16865 (74.19) | 8229 (36.18) | 0.460 | 0.754 |
| **Response 1** | 10520 (46.28%) | 0.506 | 0.517 | 7623 (72.46) | 3763 (35.77) | 0.453 | 0.747 |
| **Response 2** | 6195 (27.25%) | 0.539 | 0.552 | 4704 (75.93) | 2316 (37.38) | 0.478 | 0.753 |
| **Response 3** | 3535 (15.55%) | 0.563 | 0.568 | 2674 (75.64) | 1304 (36.89) | 0.476 | 0.747 |
| **Response 4** | 1595 (7.02%) | 0.525 | 0.515 | 1190 (74.61) | 535 (33.54) | 0.421 | 0.715 |
| **Response 5** | 646 (2.84%) | 0.539 | 0.543 | 484 (74.92) | 216 (33.44) | 0.434 | 0.675 |
| **Response 6** | 177 (0.78%) | 0.665 | 0.660 | 138 (77.97) | 65 (36.72) | 0.545 | 0.664 |
| **Response 7** | 46 (0.20%) | 0.707 | 0.654 | 39 (84.78) | 17 (36.96) | 0.570 | 0.579 |
| **Response 8** | 14 (0.06%) | 0.687 | 0.569 | 10 (71.43) | 6 (42.86) | 0.446 | 0.897 |
| **Response 9** | 4 (0.02%) | - | - | 3 (75.00) | 3 (75.00) | - | - |

Although sample size diminishes with increasing visits, concordance between the PSI and SRH remains stable, even in the cells with smaller sample size. For graphic representation (Figure 1 and Figure 2), we limited our data to 1-3 visits for visual clarity.

**Figure 1.**
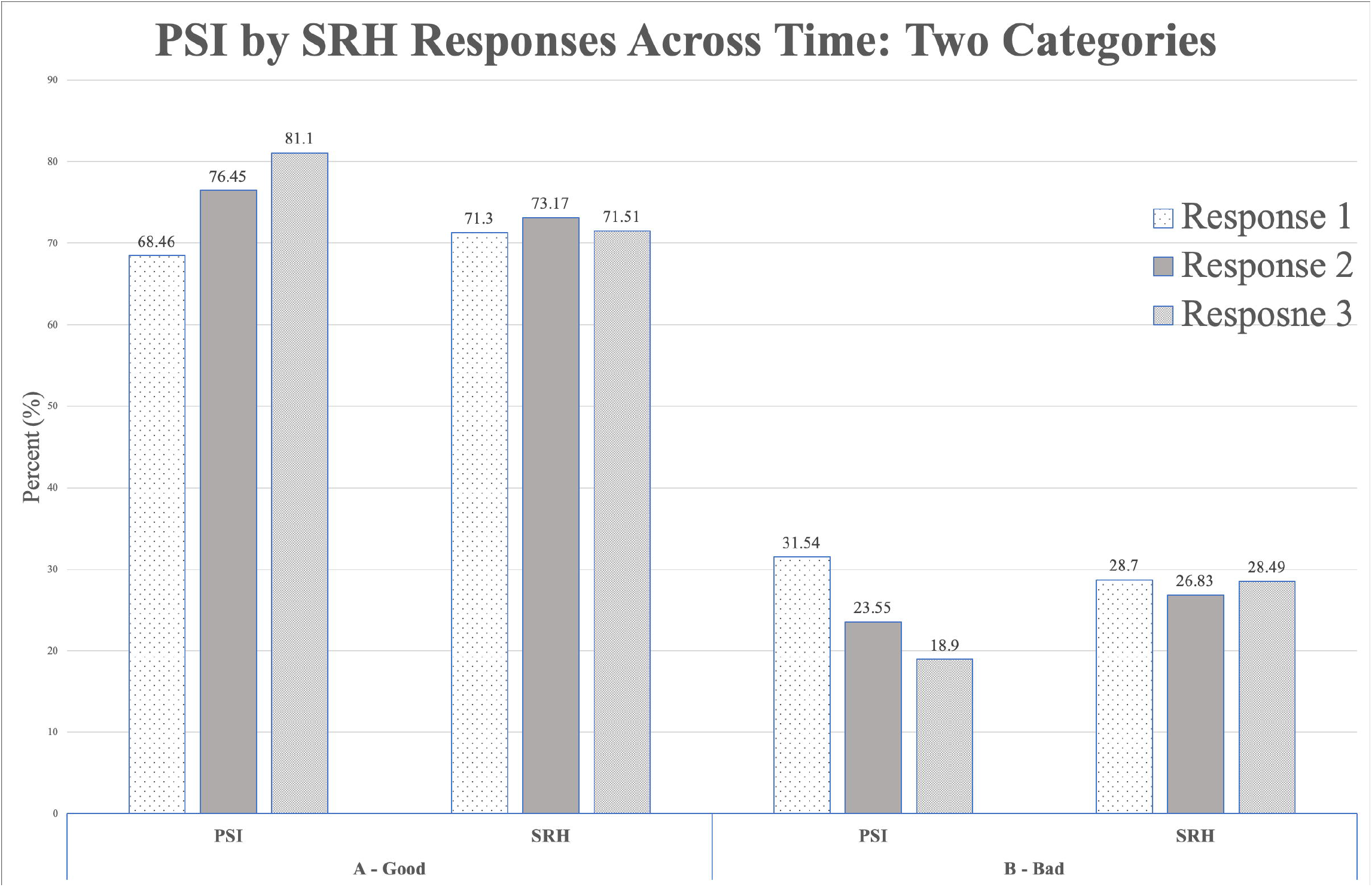

**Figure 2.**
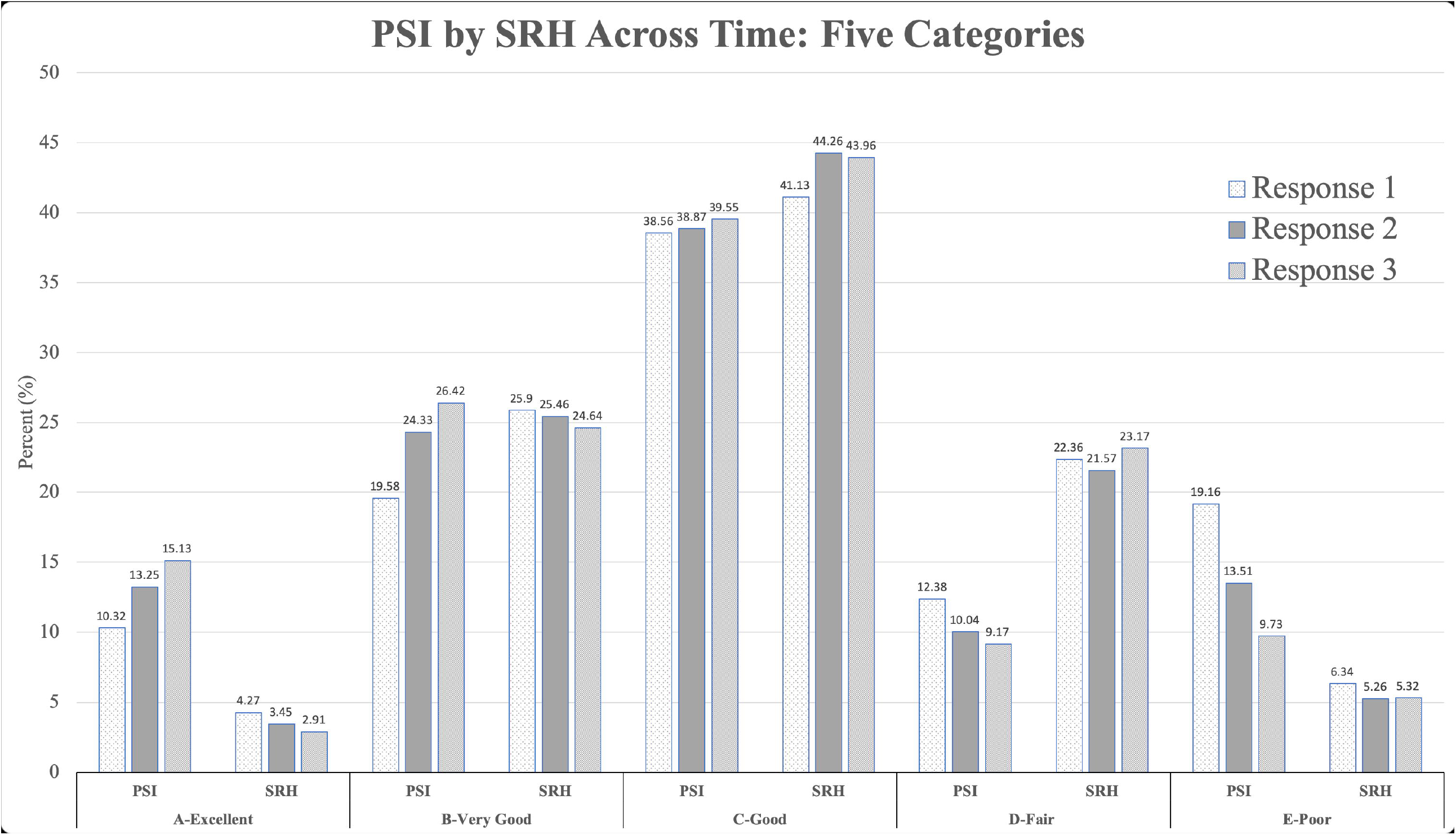

## Discussion

This study investigated the validity of the PSI, a digital first e-PROM by comparing it to the SRH in a large adult population. We found moderate association and agreement (e.g., relative concordance) between the PSI and SRH. When the PSI was scored as ordinal, it did not perfectly match the five health categories in the SRH; however, they were consistent in terms of their position as good health (Excellent, Very Good, Good) versus bad health (Fair, Poor). In other words, the PSI and SRH generally point in the same directions for self-reported health categorization.

Various analyses were performed to explore association and agreement. First, we analyzed whether collapsing PSI scores into ordinal categorical variables (versus continuous) would change the association with SRH. The results were similar between continuous (polyserial correlation) and categorical (polychoric correlation) when compared to the categorical SRH. T-tests showed no significant difference between these correlations. We also explored whether agreement between PSI and SRH were different between the first patient visit versus subsequent visits. Agreement between PSI and SRH for patients with repeated assessments remained consistent over time, suggesting consistency for the PSI from first visit to follow-up visits.

We noted that patients tended to report better health on the PSI than the SRH. In the current study, the SRH question is asked at the end of the PSI. It is possible that while answering the PSI questions, patients were reminded of their health symptoms leading them to be more likely to rate their health poorly in the SRH. The order of administration may play a role in the agreement level. Future A/B testing will explore whether the order of administration impacts the self-reported perception of well-being.

Beyond the effects of administration order, there are conceptual differences between the PSI and SRH that may contribute to the degree of agreement. While they are conceptually similar, the PSI captures more information than the SRH. We would expect a general trend of agreement or relative concordance between the two, but not to such a high degree that it would match perfectly (literal concordance).

While the focus of this study was not to assess or describe longitudinal changes between the PSI and SRH, we did observe the PSI captured improvement in symptoms over time with treatment (Table 5). In comparison, the SRH remained relatively static over responses one through three. This implies that the PSI, with its greater degree of granularity, can capture symptom changes in a way that we would not expect from a single-item question like the SRH. Further research should compare the PSI to additional tools to understand the change observed across time.

In designing a study to determine PSI validity, we found a lack of conceptually relevant tools against which to compare the PSI. Other PROMs like the PROMIS^26,27^, SF-36^28,,29^ and the Medical Symptom Toxicity Questionnaire (MSQ)^30^ are powerful assessment tools in their own right, but none were created to be digital-first in this new era of telehealth-centric care delivery. This gap in available PROMs drove us to create the PSI, and why we chose to validate it against the SRH. These results suggest that further validation of the PSI would benefit from comparing it to a PROM with similar granularity (e.g., bodily system level), even if that PROM is not a perfect conceptual match. These PROMs are similar enough that we aim to compare these tools to the PSI to better understand the PSI as a conceptually valid yet distinctly useful tool.

## Conclusion

This study showed that the PSI is a valid e-PROM that can be utilized in telehealth settings. Further validation studies should compare the PSI to PROMs of similar scope. As telehealth will inevitably continue to grow, PROMs will be increasingly used and built as exclusively digital tools. Therefore, PROMs being used in the digital space must be researched and validated within the telehealth environment. This is a paradigm shift in the world of PROM development and validation. As this field evolves we will need to assess what it means to validate a tool that is no longer administered to a captive audience in a physical waiting room, but rather, one that is engaged with virtually. Measures of engagement and ‘stickiness’ will need to be considered as we build tools that can be completed anywhere and at any time of the day. While these digital PROMs will need to be validated against previously validated tools, they must also stand up to the test of our modern, all access world.

## Data Availability

All data produced in the present study are available upon reasonable request to the authors

## Acknowledgments

The authors would like to thank Dylan Ray, Elena Céspedes, Patrick Hanaway, and Susan Silva for their support in this study.

## Authors Contribution

H. Williams, S. Steinberg, R. Berzin, and H. Hagg contributed to the conception of the study design, article preparation, and data collection. K. Leon and C. O’Shea contributed to the article preparation. All authors read and approved the final article. This statement confirms that this article has been submitted solely to this journal and is not published, in press, or submitted elsewhere.

## Author(s’) disclosure

All authors are either employees or consultants to Parsley Health at the time of analysis. All authors declare no other competing interests.

## Funding statement

This material was fully supported by Parsley Health. The funder had the following involvement with the study: study design, research, and preparation of the manuscript.

## Notes

### Competing Interest Statement

The authors have declared no competing interest.

### Funding Statement

This study did not receive any funding

### Author Declarations

The ethics committee/IRB of Stony Brook University waived and considered this study exempt from 45 Code of Federal Regulations requirements (IRB2020-00429)

## References

1. Fair Health Study analyzes telehealth. FAIR Health. https://www.fairhealth.org/article/fair-health-study-analyzes-telehealth. Published August 15, 2019. Accessed February 3, 2022.

2. Peden CJ, Mohan S, Pagán V. Telemedicine and COVID-19: an Observational Study of Rapid Scale Up in a US Academic Medical System. J Gen Intern Med. 2020;35(9):2823–2825. doi:10.1007/s11606-020-05917-9

3. Koonin LM, Hoots B, Tsang CA, et al. Trends in the Use of Telehealth During the Emergence of the COVID-19 Pandemic — United States, January–March 2020. MMWR Morb Mortal Wkly Rep 2020;69:1595–1599. DOI: http://dx.doi.org/10.15585/mmwr.mm6943a3externalicon

4. Barbosa W, Zhou K, Waddell E, Myers T, Dorsey ER. Improving access to care: Telemedicine across medical domains. Annual Review of Public Health. 2021;42(1):463–481. doi:10.1146/annurev-publhealth-090519-093711

5. Snoswell C, Taylor M, Comans T, Smith A, Gray L, Caffery L. Determining if Telehealth Can Reduce Health System Costs: Scoping Review. J Med Internet Res. 2020;22(10):e17298. URL: https://www.jmir.org/2020/10/e17298. DOI: 10.2196/17298

6. Buttorff C, Ruder T, Bauman M. Multiple chronic conditions in America. RAND Corporation. https://www.rand.org/pubs/tools/TL221.html. Published May 26, 2017. Accessed February 3, 2022.

7. Perez J, Niburski K, Stoopler M, Ingelmo P. Telehealth and chronic pain management from rapid adaptation to long-term implementation in pain medicine: A narrative review. Pain Rep. 2021;6(1):e912. Published 2021 Mar 9. doi:10.1097/PR9.0000000000000912

8. Theodore BR, Whittington J, Towle C, et al. Transaction cost analysis of in-clinic versus telehealth consultations for chronic pain: preliminary evidence for rapid and affordable access to interdisciplinary collaborative consultation. Pain Med. 2015;16(6):1045–1056. doi:10.1111/pme.12688

9. Wicklund, E Hospital’s Telehealth Program Reduces ER Visits, Treatment Costs. mHealthIntelligence.https://mhealthintelligence.com/news/hospitals-telehealth-program-reduces-er-visits-treatment-costs. Published January 25, 2019. Accessed February 10, 2022.

10. Greenhalgh, J., Gooding, K., Gibbons, E. et al. How do patient reported outcome measures (PROMs) support clinician-patient communication and patient care? A realist synthesis. J Patient Rep Outcomes 2, 42 (2018). https://doi.org/10.1186/s41687-018-0061-6

11. Chung AE, Basch EM. Incorporating the patient’s voice into electronic health records through patient-reported outcomes as the “review of systems”. J Am Med Inform Assoc. 2015;22(4):914–916. doi:10.1093/jamia/ocu007

12. Franklin P, Chenok K, Lavalee D, et al. Framework To Guide The Collection And Use Of Patient-Reported Outcome Measures In The Learning Healthcare System. EGEMS (Wash DC). 2017;5(1):17. Published 2017 Sep 4. doi:10.5334/egems.227

13. Williams H, Steinberg S, Berzin R. The Development of a Digital Patient-Reported Outcome Measurement for Adults With Chronic Disease (The Parsley Symptom Index): Prospective Cohort Study. JMIR Form Res. 2021;5(6):e29122 DOI: 10.2196/29122

14. Cullati S, Mukhopadhyay S, Sieber S, et al. Is the single self-rated health item reliable in India? A construct validity study. BMJ Global Health. 2018;3:e000856.

15. Schnittker J, Bacak V. The increasing predictive validity of self-rated health. PLoS One. 2014;9(1):e84933. Published 2014 Jan 22. doi:10.1371/journal.pone.0084933.

16. Kaplan MS, Berthelot JM, Feeny D, McFarland BH, Khan S, Orpana H. The predictive validity of health-related quality of life measures: mortality in a longitudinal population-based study. Qual Life Res. 2007;16(9):1539–1546. doi:10.1007/s11136-007-9256-7

17. U.S. Department of Health and Human Services. Protection of human subjects. Code Fed Regul Public Welfare. 1995;Title 45(Sections 46-101 to 46-409).

18. Patient-Reported Outcome Measures: Use in Medical Product Development to Support Labeling Claims. Federal Drug Administration (FDA) 2019. https://www.fda.gov/regulatory-information/search-fda-guidance-documents/patient-reported-outcome-measures-use-medical-product-development-support-labeling-claims.

19. Zajacova A, Dowd JB. Reliability of self-rated health in US adults. American Journal of Epidemiology. 2011;174(8):977–983. doi:10.1093/aje/kwr204

20. Ranganathan P, Pramesh CS, Aggarwal R. Common pitfalls in statistical analysis: Measures of agreement. Perspect Clin Res. 2017;8(4):187–191. doi:10.4103/picr.PICR_123_17

21. Julius Sim, Chris C Wright, The Kappa Statistic in Reliability Studies: Use, Interpretation, and Sample Size Requirements. Physical Therapy, Volume 85, Issue 3, 1 March 2005, Pages 257–268, https://doi.org/10.1093/ptj/85.3.257

22. Jürges H, Avendano M, Mackenbach JP, Are different measures of self-rated health comparable? An assessment in five European countries. Eur J Epidemiol. 2008;23(12):773–781. doi:10.1007/s10654-008-9287-6

23. P.F. Watson, A. Petrie, Method agreement analysis: A review of correct methodology, Theriogenology, Volume 73, Issue 9, 2010, Pages 1167-1179, ISSN 0093-691X, https://doi.org/10.1016/j.theriogenology.2010.01.003.

24. Zaki R, Bulgiba A, Ismail R, Ismail NA. Statistical methods used to test for agreement of medical instruments measuring continuous variables in method comparison studies: a systematic review. PLoS One. 2012;7(5):e37908. doi:10.1371/journal.pone.0037908

25. R: A language and environment for statistical computing. R Foundation for Statistical Computing. [2016-11-01]. https://www.R-project.org/

26. Cella D, Riley W, Stone A, et al. The Patient-Reported Outcomes Measurement Information System (PROMIS) developed and tested its first wave of adult self-reported health outcome item banks: 2005-2008. J Clin Epidemiol. 2010;63(11):1179–1194. doi:10.1016/j.jclinepi.2010.04.011

27. DeWalt DA, Rothrock N, Yount S, Stone AA; PROMIS Cooperative Group. Evaluation of item candidates: the PROMIS qualitative item review. Med Care. 2007;45(5 Suppl 1):S12–S21. doi:10.1097/01.mlr.0000254567.79743.e2

28. LoMartire R, Äng BO, Gerdle B, Vixner L. Psychometric properties of Short Form-36 Health Survey, EuroQol 5-dimensions, and Hospital Anxiety and Depression Scale in patients with chronic pain. Pain. 2020;161(1):83–95. doi:10.1097/j.pain.0000000000001700

29. Cordier R, Brown T, Clemson L, Byles J. Evaluating the Longitudinal Item and Category Stability of the SF-36 Full and Summary Scales Using Rasch Analysis. Biomed Res Int. 2018;2018:1013453. Published 2018 Nov 4. doi:10.1155/2018/1013453

30. MSQ-Medical Symptom/Toxicity Questionnaire. Hyman, MA. [2020-05-01]. http://drhyman.com/downloads/MSQ_Fillable.pdf.

